# Reassessing referred sensations following peripheral deafferentation and the role of cortical reorganisation

**DOI:** 10.1101/2021.12.08.21267128

**Authors:** Elena Amoruso, Devin B. Terhune, Maria Kromm, Stephen Kirker, Dollyane Muret, Tamar R. Makin

**Affiliations:** Institute of Cognitive Neuroscience, University College London, London WC1N 3AZ, UK; Department of Psychology, Goldsmiths, University of London, London SE14 6NW, UK; Cambridge University Hospitals NHS Foundation Trust, Cambridge CB2 0QQ, UK; Wellcome Trust Centre for Neuroimaging, University College London, London WC1N 3AR, UK

## Abstract

**Background and Objectives:** Tactile sensations referred to body parts other than those stimulated have been repeatedly described across a wide range of deafferentation and neuropathic pain conditions, including amputation, complex regional pain syndrome, spinal cord injury, and brachial plexus avulsion. Common to all interpretations of referred sensations is the notion that they result from central nervous system (CNS) reorganisation. For example, in amputees, sensations referred to the phantom limb following touches on the face have been classically interpreted as the perceptual correlate of cortical remapping of the face into the neighbouring missing-hand territory in primary somatosensory cortex (S1). Here, using the prominent model of acquired upper-limb amputation, we investigated whether referred sensations reports are associated with cortical remapping or can instead be attributed to demand characteristics (e.g., compliance, expectation, and suggestion), which have been shown to greatly influence self-reports of bodily sensations and were uncontrolled in previous assessments.

**Methods:** Unilateral upper-limb amputees (N=18), congenital one-handers (N=19), and two-handers (N=20) were repeatedly stimulated with PC-controlled vibrations on ten body-parts and asked to report on each trial the occurrence of any concurrent sensations on their hand(s). To further manipulate expectations, we gave participants the suggestion that some of these vibrations had a higher probability to evoke referred sensations. To evaluate remapping, we analysed fMRI data in S1 from two tasks involving movement of facial and whole-body parts, using univariate and multivariate approaches.

**Results:** The frequency and distribution of reported referred sensations were similar across groups, with higher frequencies in the high expectancy condition. In amputees, referred sensations were evoked by stimulation of multiple body-parts and reported in both the intact and phantom hand. The group profiles for referred sensations reports were not consistent with the observed patterns of S1 remapping.

**Discussion:** These findings weaken the interpretation of referred sensations as a perceptual consequence of post-deafferentation CNS reorganisation and reveal the need to account for demand characteristics when evaluating self-reports of anomalous perceptual phenomena for both research and clinical assessments purposes.

## INTRODUCTION

Most amputees experience phantom sensations from their missing limb, typically described as itching, tingling, or numbness [1]. These sensations typically manifest spontaneously but can sometimes also be triggered through stimulation of another body-part. Most commonly, phantom sensations can be evoked by touch applied to the residual limb (stump). This is believed to reflect peripheral reinnervation, where the severed sensory nerves, initially targeting e.g., the hand, reinnervate the surrounding tissue [2]. A more curious example of evoked phantom sensations in upper-limb amputees comes from anecdotal reports that touching the face (e.g., while shaving) can elicit tingling sensations on the phantom hand. In a famous series of case studies [3–6], a small group of patients reported experiencing referred sensations from the ipsilateral face to the phantom hand. In some cases, the referred sensations were modality specific, with, for example, hot water applied to the face eliciting a warm sensation on the phantom hand. Strikingly, in most of these patients, neighbouring sites on the face elicited sensations on neighbouring fingers, suggesting a shared topographical organisation of the face and phantom hand.

Phantom referred sensations evoked by facial stimulation have been commonly interpreted as the perceptual correlate of primary somatosensory cortex (S1) remapping, observed in classic electrophysiological studies in non-human primates [7]. Specifically, it was shown that, following long-term arm deafferentation, the deafferented hand territory became responsive to touch applied on the monkey’s chin. It was therefore suggested that following a similar remapping process in humans, face-induced activity in the missing hand area will be perceived as arising from the missing hand (hereafter, ‘the perceptual remapping hypothesis’) [4,5,8,9]. This theory has been further extended to consider the neural origins of phantom limb pain [10], as well as analogous referred sensations described in other neurological disorders [11–14].

However, subsequent studies using more systematic stimulation paradigms found that phantom referred sensations could be evoked by touches on various body-parts. This includes body-parts that have not been considered to invade the missing hand cortical area, such as the feet, trunk, and neck, in some cases even contralateral to the missing hand [3,15–19]. These reports, as well as recent fMRI studies that dispute the existence of a large-scale S1 facial remapping post-amputation in humans [20–23], challenge the perceptual remapping hypothesis.

An alternative mechanistic framework of referred sensations is that previous studies, lacking adequate controls, might have been confounded by the demand characteristic which are typical of experimental settings where the desired outcome (or response) is known or can be implicitly inferred by the context [24]. Beyond compliance effects, demand characteristics conveyed by a procedure or intervention can also produce genuine experiences. For example, verbal suggestions can elicit robust changes in perceptual states [25,26]. It is increasingly recognised that demand characteristics in various forms may function as confounds in a variety of experimental paradigms [27–31]. Therefore, rather than cortical remapping, reports of referred sensations in previous experiments might be driven by both explicit suggestions from experimenters (e.g., “this procedure will produce this experience”) or implicit cues that promote expectations for specific experiences.

Here we used vibrotactile stimulation (used to evoke reliable referred sensations in previous reports, e.g.,[15,17,18]) of ten body-parts in a group of upper-limb amputees experiencing spontaneous phantom sensations. We also tested two control groups who do not report feelings of phantom sensations: two-handed individuals and individuals born with one hand. We manipulated participants’ expectations for referred sensations with explicit verbal suggestions that specific vibro-tactile stimuli were more likely to evoke referred sensations on their hands (and even if that hand is missing). To evaluate whether referred sensations are associated with cortical remapping, we analysed functional neuroimaging (fMRI) data, collected from two sensorimotor tasks in the same participants, using both univariate and multivariate approaches. We find that referred sensations can be generated and influenced by demand characteristics but no evidence to support the perceptual remapping hypothesis.

## METHODS

### Participants

Eighteen individuals with acquired unilateral upper-limb amputation (hereafter Amputees; mean age = 52 ± 12.2 (SD) y/o, 6 women, 10 missing the right upper-limb; see Table 1 for details about amputation, phantom limb pain and sensations), 19 individuals with congenital unilateral transverse arrest (hereafter One-handers; mean age = 44 ± 14.3 (SD) y/o, 11 women, 7 missing right upper-limb), and 22 two-handed individuals (hereafter Two-handers; mean age = 45.5 ± 9.5 (SD) y/o, 10 females, 6 left-handed) were tested (see Supplementary section for further details). The study was designed in accordance with the Declaration of Helsinki and was approved by the UK Health Research Authority (HRA) and Health and Care Research Wales (HCRW) committee (18/LO/0474). All participants provided written informed consent prior to participating in the study.

**Table 1.**
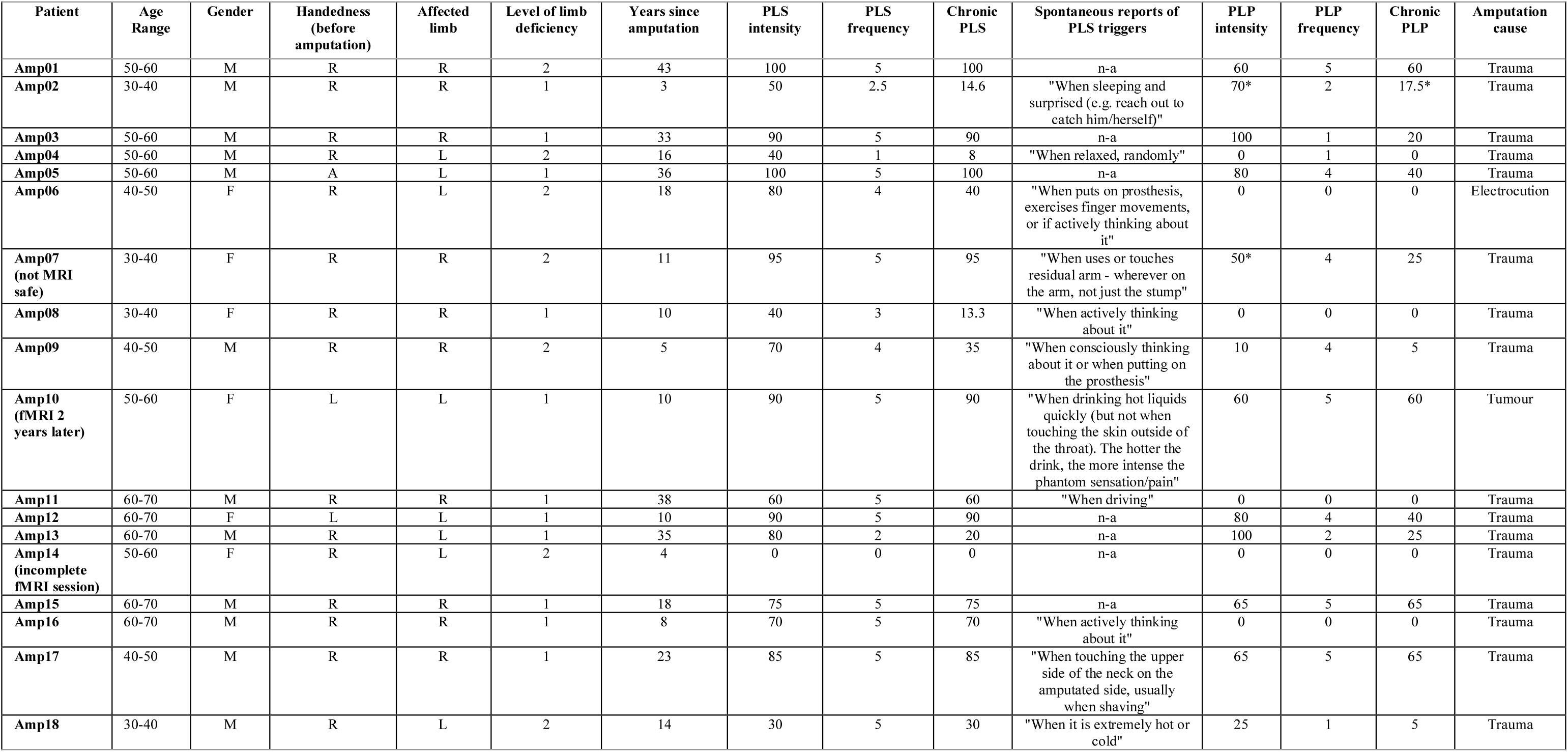
Demographic details for Amputees. Level of limb deficiency: 1= above elbow (transhumeral), 2= below elbow (transradial); L= left, R= right. PLS= phantom limb sensations; PLP= phantom limb pain. PLS & PLP frequency: 0= no sensation or pain, 1= once or less per month, 2= several times per month, 3= once a week, 4= daily, 5= all the time. PLP intensity: worst PLP experience during the last week or in a typical week involving PLP, with 0= no pain, 100= worst pain imaginable. A chronic measure of PLP for correlation analyses was calculated by dividing the worst PLP intensity in the last week by PLP frequency. Spontaneous reports of PLS triggers: responses to the item: “Does your phantom sensation “come to life” following specific actions or situations (for example, putting on your prosthesis, when you touch your face)?”. ^*^PLP intensity rating was on average

### Referred sensations task

#### Procedure

Participants sat in front of a computer screen (2560×1440, 60Hz) and a pedal was positioned underneath each foot for response collection. Vibrating motors (diameter: 10mm, thickness: 2.7mm, operating voltage: 5.5V DC) were secured with surgical tape on ten body-parts (Figure 1A). Participants were informed that some of the vibrations were stimulating a newly discovered type of afferent fibres that could evoke sensations not only on the stimulated body-part but also on other areas, most commonly on the hands (see Supplementary section for full instructions). Participants were told that a red or grey circle display would indicate whether an incoming stimulus had high or low probability, respectively, of evoking these dual sensations (hereafter, “High” and “Low” expectation conditions). All stimuli were, in effect, simple 500ms vibrating trains. To induce the feeling that vibrations may vary, and thus increase the belief in the suggestion, trains were delivered at an intensity well above detection threshold at three frequencies (15Hz, 12Hz, or 9Hz), with equal distribution across the different locations and cues. Participants were instructed to stay still and to focus on potential sensations arising from their hands.

**Figure 1.**
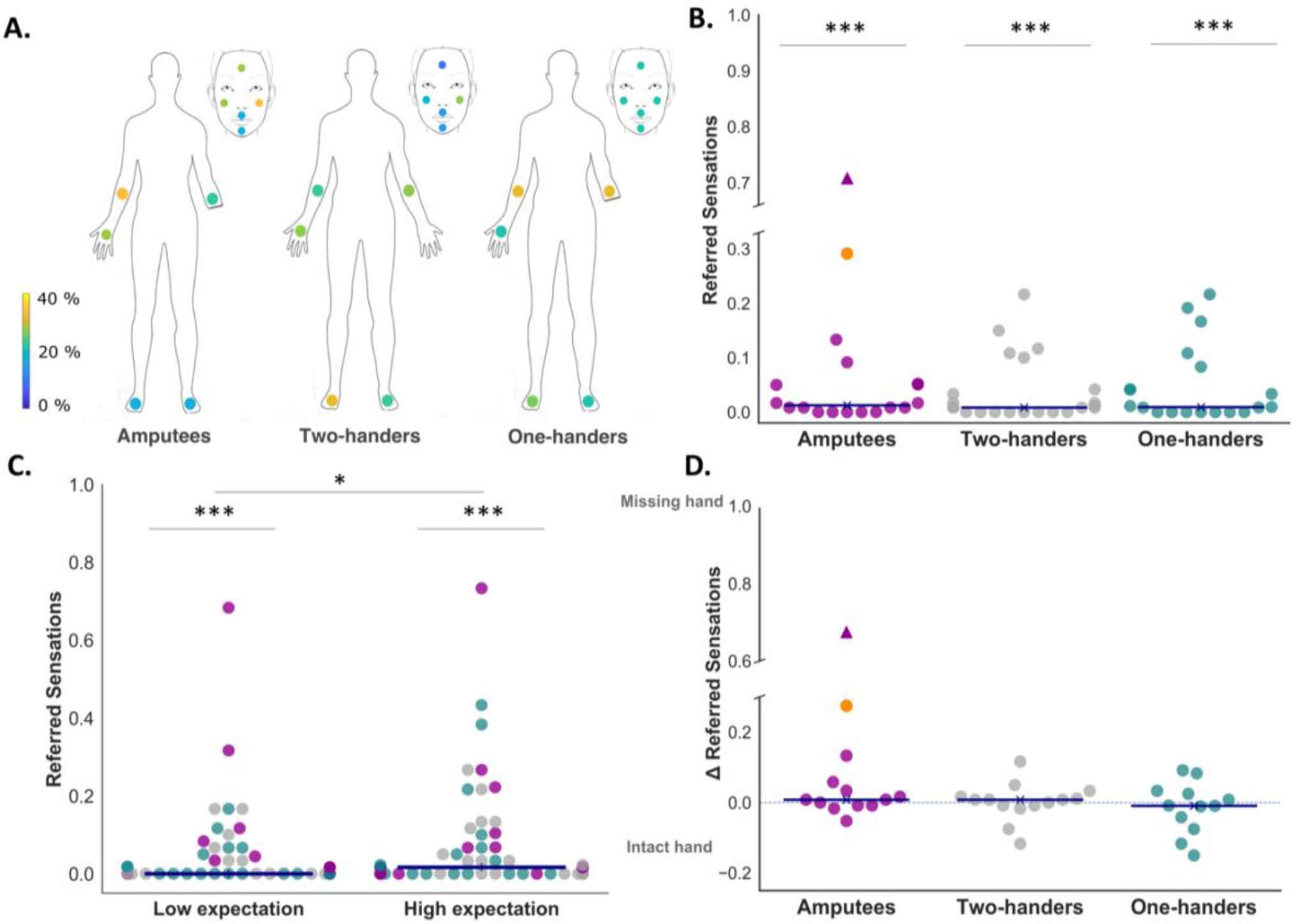
Global pattern of referred sensations across groups. **A)** Circles on the body silhouettes indicate the stimulated locations. The colour code indicates the percentage of participants in each group reporting at least one referred sensation at a given location. **B)** Proportion of referred sensations reported in each group across the stimulated locations. All groups reported referred sensations significantly above zero. **C)** Proportion of referred sensations reported across groups in the Low and High expectation conditions. (***p<.005, *p<.05). **D)** Lateralised referred sensations across all stimulated locations, calculated by subtracting the proportion of referred sensations reported on the intact/dominant hand from the proportion of responses on the phantom/missing/non-dominant hand, in Amputees, One-handers and Two-handers, respectively. Positive values reflect referred sensations reported more towards the phantom/missing/non-dominant hand. Participants reporting zero referred sensations across the experiment were excluded (33.9% of the total sample). Each dot represents one participant; horizontal blue lines represent the group medians. In B) and D) participant Amp05, who reported high rates of phantom referred sensations and who could be scanned (contrary to Amp07, triangle symbol), is highlighted in orange to ease qualitative comparison with fMRI results shown in Figure 2.

Except for the instruction phase, the experiment ran in a strictly automatised way, with the experimenter leaving the room and visual cues, stimuli and responses delivered and collected by the software (MATLAB r2017a). Each trial began with the red/grey visual cue displayed for 700ms, followed by the 500ms vibration on one body-part. Next, a question appeared on the screen: “Have you just felt one stimulus or more than one?”. Participants were instructed to press with the left foot to respond “one sensation” and with the right to respond “more than one sensation”. When reporting multiple sensations, participants were further asked to respond, with the corresponding foot pedal, to the question: “Was it on the right or the left hand?”. Amputees and One-handers were briefed that this question also related to their phantom/missing hand (respectively). The experiment included 120 trials, equally distributed across the 10 stimulated locations, 60 trials per expectation cue (High/Low), and 40 trials per vibration frequency (9Hz, 12Hz, 15Hz). Due to technical issues during data collection, two Amputees, one One-hander and one Two-hander performed a reduced number of trials (90).

#### Analyses

Trial-level data were aggregated into participant-level data by calculating the proportion of reported referred sensations in each condition. To focus on referred sensations reported on the phantom hand, while accounting for responses on the intact hand, lateralisation scores were calculated by subtracting the proportion of intact/dominant hand to phantom/missing/non-dominant hand referred sensations reports, in Amputees, One-handers and Two-handers respectively. Only participants reporting at least one referred sensation in a given condition of interest were included in this calculation. This resulted in different sample sizes across analyses/groups (see Results).

### Functional MRI tasks

#### Procedure

The scanning session was completed on the same day as the behavioural task, apart from one Amputee who was scanned only 2 years later (due to Covid-19 restrictions; Amp10 in Table 1). Full details are provided in the Supplemental Material. In brief, the conditions included in the present analysis involved visually-instructed body-part movements. For the body task, this involved one of five body-parts (12s blocks): intact/dominant hand (open/closing the fingers), residual/non-dominant arm (flexing the most distal residual joint, the elbow for Two-handers), right or left toes (wiggling the toes) or lips (puckering the lips) (Figure 2B). For the face task, this involved one of five movements (8s blocks): raise the eyebrows (i.e., forehead), flare nostrils (i.e., nose), puckering lips (i.e., lips), and flex the left or right thumb (or phantom thumb, if available). When phantom sensations were not vivid, participants were asked to imagine performing the movement. To confirm that appropriate movements were made at the instructed times, task performance was visually monitored online for both tasks. These datasets were recently used for other purposes (i.e., body task used as functional localiser in [32] and face task analysed in more detail in [21]), but the body dataset was not used before to assess remapping.

**Figure 2.**
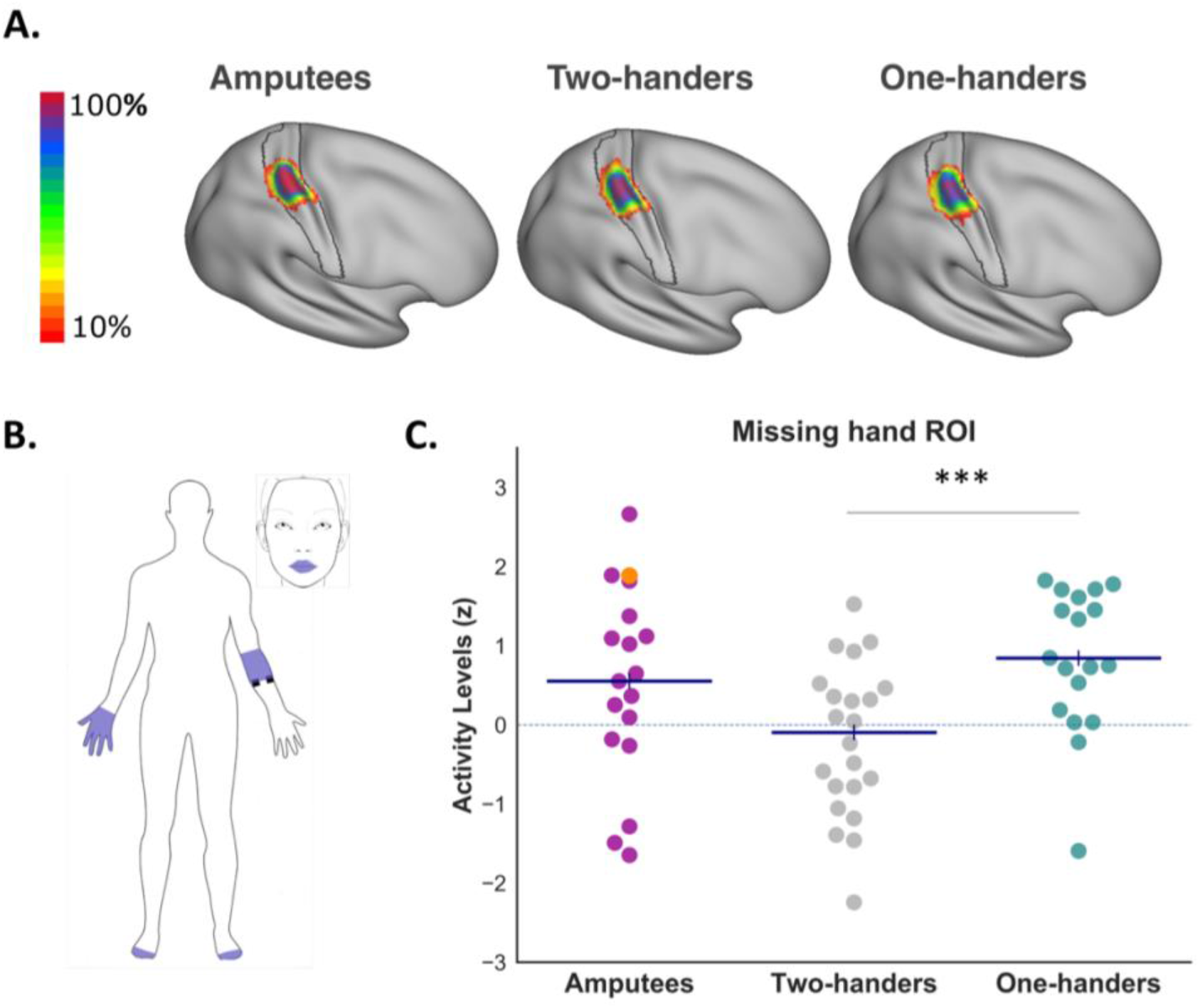
Remapping in S1 missing-hand area. **A)** Inter-participant consistency maps for the missing/non-dominant hand S1 regions of interest (ROIs) across the three groups. The colour code represents the number of participants with overlapping ROIs in standard MNI space. The black contour shows the anatomical delineation of S1 used for ROI definition, as detailed in Supplemental Material. **B)** Body-parts included in the fMRI body task. **C)** Average BOLD activity levels in the missing/non-dominant hand ROI, evoked by movement of the body-parts shown in B (***p<.005). Each dot represents one participant, horizontal blue lines represent the group medians. Note that, overall, increased remapping (though see also Figure S2) can be qualitatively observed for participant Amp05 (highlighted in orange), who reported high rates of phantom referred sensations (Figure 1).

#### Acquisition and analysis

3T MRI data acquisition and pre-processing followed standard procedures, as detailed in the Supplemental Material (see also [21,32]). Functional data was analysed in individual’s native functional space and pre-processed using FSL-FEAT (v.6.00). Voxel-wise General Linear Model (GLM) was applied to the data using FEAT to obtain statistical parametric maps for each movement. A regressor of interest for each movement was convolved with a double-gamma hemodynamic response function and its temporal derivative was included. The six motion parameters were included as regressors of no interest. A seventh regressor of no interest was used to regress out volumes with large head movements (>0.9mm). A contrast relative to rest was set up for each movement, resulting in five contrasts for each task.

Anatomical hand regions of interest (ROI) were defined bilaterally at the individual level within S1 (Brodmann areas 3a, 3b, 1 and 2 approximately 1cm below and above the hand knob; see Supplemental Material for more details). The z statistic timeseries from all voxels of each ROI obtained for each movement were extracted and averaged. For the face task, cross-validated Mahalanobis distances, extracted from a Representational Similarity Analysis (RSA)[33], were used to assess the multivariate relationship between activity patterns generated by the contralateral thumb and the four face parts, as detailed in the Supplemental Material. As One-handers do not have a representation of their missing hand, they were excluded from this analysis.

### Statistical analyses

All statistical analyses were carried out using JASP (V.0.14.1). To identify violations of the normality assumption, Shapiro-Wilk tests were run. No outliers were removed from the analyses to not exclude individuals showing potentially high referred sensations rates and/or large remapping. For non-significant comparisons of interest, Bayesian t-tests (or non-parametric equivalents) were conducted, with a Cauchy prior width set to 0.707 (default). We report Bayes Factors (BF_10_), showing the relative support for the alternative hypothesis [34]. As measure of effect size, rank-biserial correlations (r_B_) or Cohen’s d are reported. Kendall’s Tau correlations were used to investigate whether referred sensations reports or cortical remapping were related to PLP in Amputees.

For the behavioural task, as normality was consistently violated across conditions, nonparametric tests (i.e., Wilcoxon signed-rank, Mann-Whitney, and Kruskal-Wallis tests) were used to test for within- and between-subject differences in the overall proportions and lateralisation of reported referred sensations.

fMRI data was analysed using mixed ANOVAs with the between-subject factor of Group (three groups) and a repeated-measure factor of Hemisphere (intact/dominant x deprived/non-dominant), and age as a covariate to account for age differences in cortical activation. If assumptions of normality were violated, non-parametric equivalents are also reported. Post-hoc comparisons were conducted with a Bonferroni correction for multiple comparisons (*α*=.025; uncorrected p-values reported in the text). See Supplemental Material for additional analyses.

## RESULTS

### When sharing similar expectations, amputees do not report more referred sensations than one- and two-handers

First, we examined if Amputees are more prone to report referred sensations, relative to One- and Two-handers. We found that all groups reported experiences of referred sensations significantly above zero (Z≥78, p<.005, r_B_=1.0, for all groups; Figure 1B), with no differences in the proportion of trials where referred sensations were reported across stimulation frequencies (*X*^*2*^≤2.311, p≥.315 for all groups). This result demonstrates that, when given the same instructions, any sample, irrespective of amputation, can report referred sensations.

Trials in the High expectation condition evoked more reports than in the Low expectation condition (N=59, Z=206.5, p=.047, r_B_=-.38), indicating that referred sensations can be enhanced through explicit suggestions. We observed no group differences for this suggestion effect (*X*^*2*^=.560, p=.756), with Amputees’ difference (high-low) scores not different from One- and Two-handers (both U>173, p>.499, r_B_<-.124, BF_10_<.403). Interestingly, referred sensations were reported more frequently relative to zero across all groups even during the Low expectation condition (N=59, Z=351, p≤.001, r_B_=1.0; 42.4% of participants reported >0 referred sensations), indicating that our experimental setup educed robust demand characteristics effects even when expectations were low. For this reason, further analyses were based on the proportion of referred sensations collapsed across expectancy conditions. Crucially, the overall proportion of reported referred sensations did not differ across groups (*X*^*2*^=.338, p=.845), and Amputees (N=18) did not report more referred sensations than One-(N=19) or Two-handers (N=22) (both U<220, p>.550, rB<.111, BF10<.355). In summary, participants of all groups responded positively, but similarly, to the suggestion/expectation cues, regardless of amputation.

### Sensations are not differentially referred to amputees’ phantom hand

Next, we assessed the specificity of Amputees’ phantom hand as a target for referred sensations. Both One-(N=12) and Two-handers (N=14) reported similar proportions of referred sensations across the two hands (both Z≤53.5, p≥.583, r_B_≤-.192, BF_10_≤.353). If referred sensations result from deprivation-triggered cortical remapping, then they should occur more frequently on Amputees’ phantom hand and scale with remapping measures in the missing-hand region. However, we found no significant differences in the proportion of reported sensations across the two hands in Amputees (N=13; Z=59, p=.125, r_B_=.513, BF_10_=1.11), or in the lateralisation towards the missing/non-dominant hand across groups (*X*^*2*^=2.498, p=.287). Amputees did not show greater lateralisation towards the phantom hand than One- or Two-handers (both U≥106, p≥.134, r_B_≤.359, BF_10_≤.940), indicating that they were not more inclined to report referred sensations on the phantom hand than One- and Two-handers on their missing/non-dominant hand, respectively. Finally, no differences were observed between One-handers’ and Two-handers’ lateralisation scores (U=70, p=.486, r_B_=-.167, BF_10_=.417).

To gain further insights into the hypothesized link between referred sensations and S1 remapping, we assessed average activity levels evoked by movement of multiple body-parts in the hand area of each hemisphere. Compared to Two-handers (N=22), increased activity levels (i.e., remapping) were found in the missing-hand area of One-handers (N=19) (U=86, p<.001, r_B_=-.589), with only a marginal difference between Two-handers and Amputees (N=17) (t_(37)_=-1.928, p=.062, d=-.623, BF_10_=1.315), resulting in a significant interaction between Groups and Hemispheres (F_(2,54)_=8.753, p<.001, *η*^2^=.039; non-parametric equivalent: *X*^2^=10.497, p=.005; Figure 2C; see Figure S1 for data in the intact/dominant hand area). No difference in activity levels in the deprived hemisphere was found between Amputees and One-handers (U=198, p=.257, r_B_=.226, BF_10_=.506) (see Figure S2 for a breakdown of the body-parts showing increased activity in the missing-hand area). These group differences are qualitatively, though clearly, distinguishable from the even inter-group profile observed for the referred sensations reports. In other words, the non-significant group difference for reported sensations on the phantom hand is unlikely to be due to lack of remapping (see Supplemental Material for a replication of these and subsequent results using samples matched to the referred sensation task analyses).

Given the relation previously reported between the degree of sensorimotor remapping and PLP,[10,35](although see also [36]) we also explored the correlation between the propensity to report referred sensations on the phantom hand (while accounting for intact hand reports) and chronic PLP (Table 1), and found no significant correlation (N=18, r_Tau_=.074, p=.692, BF_10_=.327). In line with this, no significant correlation was found between fMRI activity levels in the missing-hand area and chronic PLP in Amputees (N=17, r_Tau_=.103, p=.582, BF_10_=.360).

### Face-evoked referred phantom sensations are not reflected in shared S1 representation

According to the “perceptual remapping hypothesis”,[3–5,8] phantom hand referred sensations in amputees should more likely occur following face stimulation, due to the proximity in S1 representations. In accordance with this hypothesis, we found that face-evoked referred sensations were reported marginally more frequently on the phantom/missing/non-dominant rather than on the intact/dominant hand in Amputees (N=10) (Z=46.5, p=.058, r_B_=.691, BF_10_=2.215), but not in One-handers (N=9) and Two-handers (N=10) (both Z≤30, p≥.836, r_B_≤.091, BF_10_≤.330) (Figure 3). However, no significant group difference emerged (*X*^*2*^=3.417, p=.181), with Amputees’ phantom lateralised face-evoked referred sensations not significantly different from One-Handers’ (both U≤69.5, p≥.109, r_B_≤444, BF_10_≤.952), and no significant differences between One-handers and Two-handers (U=41.5, p=.805, r_B_=-.078, BF_10_=.414), which could be attributed to reduced statistical power, as indicated by the Bayes factors (see also Figure S3 for analyses of referred sensations evoked by other stimulation sites).

**Figure 3.**
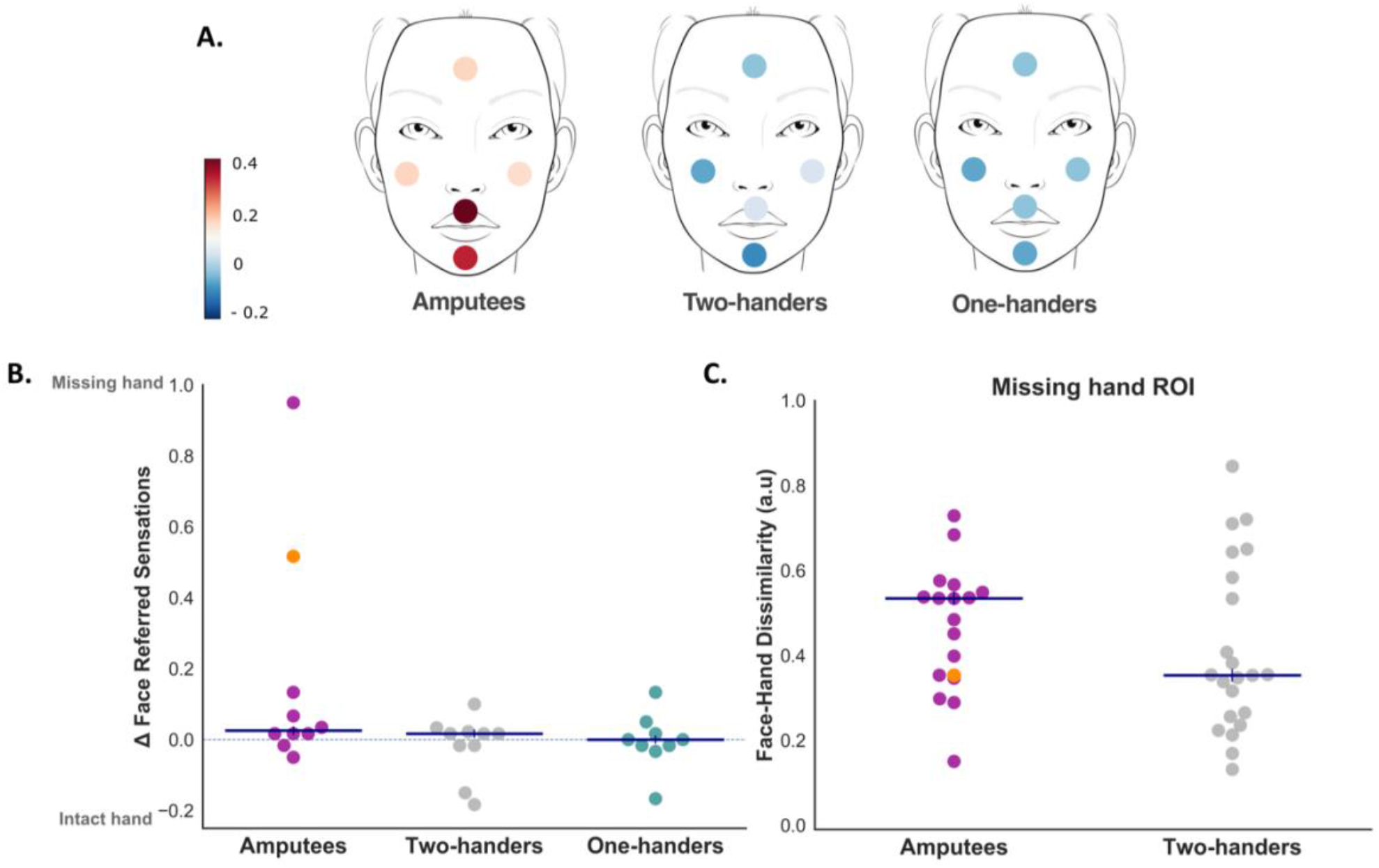
Face-evoked referred sensations and face-hand representational content in the missing-hand area. **A)** The colour code indicates the group median lateralisation scores of referred sensations from each of the stimulated face locations. **B)** Lateralised referred sensations across the five face locations (60 trials). Participants reporting zero referred sensations on any hand across the face trials are excluded (50.8% of total sample). **C)** Multivariate representational dissimilarity between activity patterns evoked in the missing/non-dominant hand ROI by facial (lips, nose, forehead) and contralateral thumb movements in Amputees and Two-handers (One-handers excluded as they cannot perform phantom movements). Whole samples are included, each dot represents one participant, horizontal blue lines represent group medians. Participant Amp05 is highlighted in orange: despite reporting high rates of referred sensations, no decreased dissimilarity between face and phantom hand representations is observed.

To provide more direct insight into the relationship between face and phantom hand representation, multivariate representational similarity analysis (RSA) was used (Figure 3C; see also Figure S2 for univariate results). Comparing the representational dissimilarity between activity patterns evoked by face and contralateral thumb movements in the hand area revealed no significant differences between Amputees and Two-handers (F_(1,36)_=.372, p=.546, *η*^2^=.009), and no interaction with the Hemisphere (F_(1,36)_=1.402, p=.244, *η*^2^=.004). Follow-up comparisons revealed no significant difference between Hemispheres in Amputees (t_(16)_=.619, p=.545, d=.150, BF_10_=.295), as well as no group difference in the dissimilarities observed in the missing/non-dominant hand area (t_(37)_=-1.047, p=.302, d=-.338, BF_10_=.483), with Amputees showing, if anything, more face-hand dissimilarity than Two-handers (Figure 3C). In other words, we did not find any evidence for shared information content between the phantom hand and the face in the missing hand area (see also [21], for a further characterisation of the lack of facial remapping in this cohort of patients).

## DISCUSSION

Here, we studied referred sensation reports in a group of upper-limb amputees experiencing spontaneous phantom limb sensations, in comparison to two control groups who had not undergone amputation (congenital one-handers and two-handers). Amputees did not report more induced referred sensations than the control groups. In all groups, referred sensations could be evoked from stimulation of multiple body-parts on both sides of the body, irrespective of inter-group differences in S1 remapping (Figure 2, S2; see also [19,37]). When sensations were evoked by face stimulation, we found marginal evidence for amputees differentially referring these sensations to the phantom hand, though this did not drive a significant interaction across groups. Importantly, this marginal effect in amputees is likely not consequential to shared S1 processing between the face and the phantom hand in the missing-hand territory (Figure 3; see also [20–23] for evidence of minimal face remapping). Overall, our findings contrast with the mainstream hypothesis that referred sensations are the perceptual correlate of post-amputation S1 remapping [4,5,8,9]. The lack of consistency with measures of cortical remapping and the fact that referred sensations were reported not only on amputees’ phantom hand but also by the control groups, call for a general reassessment of this phenomenon and its neural bases. Specifically, while referred sensations might be genuinely and spontaneously experienced by some amputees, the experimental methods used to date to assess this phenomenon clearly contain demand characteristics that at best will contaminate any true effects.

It is well recognized that self-reported phenomena are particularly susceptible to the confounding impact of demand characteristics [27,38–40], yet there have been no attempts to control for these effects in referred sensations testing paradigms. We demonstrate that self-reported referred sensations can be triggered by experimental settings. This is well evidenced not only by the fact that the control groups reported referred sensations in the first place, but also, more directly, by a greater tendency across groups to report referred sensations when they were given the suggestion that these sensations were more likely to occur (‘High’ expectation condition). Expectancy-mediated changes in self-reported experiences can be driven by simple compliance or genuine changes in perception. Although our data does not allow us to dissociate genuine perceptual changes from compliance effects, the observation that congenital one-handers reported referred sensations also on their missing hand, on which touch had never been experienced, and thus no sensations can truly be referred to, suggests that behavioural compliance may have played a prominent role in our results.

The lack of control for demand characteristics poses serious limitations to the interpretation of previous accounts of referred sensations, as well as of other related phenomena that solely rely on self-assessed outcomes without accounting for both experimenter and participants’ expectation (e.g., PLP treatment)[41]. It is important to note that our findings do not rule out the spontaneous occurrence of referred sensations in selected amputees. Indeed, one participant reported experiencing referred sensations from the neck in his daily life (Amp17 in Table 1) and another (Amp05) reported a “classical” pattern of face-elicited referred sensations in our study (though this did not translate in increased cortical face remapping, as highlighted in Figure 3, S2). However, our findings clearly show that this phenomenon can be induced by suggestion and expectation. Insofar as referred sensations reports were plausibly driven, or augmented, by such confounds in previous research, they cannot provide a solid perceptual foundation for theories about functional brain reorganisation [9] or for novel efforts to create ecological tactile feedback interfaces for prosthetic limbs [9,42,43]. We hope that our findings will promote greater consideration of experimental demand characteristics in future research on other neurological phenomena.

## Data Availability

Anonymised data will be made available prior to publication in a public depository (https://osf.io).

## Acknowledgments

This work was supported by an ERC Starting Grant (715022 EmbodiedTech) and a Wellcome Trust Senior Research Fellowship (215575/Z/19/Z), awarded to TRM. DBT is supported by the Gyllenbergs Foundation. We thank Benjamin Kop for contributing to data collection and pre-processing. We thank Opcare and Arabella Bouzigues for their help with participants recruitment, and our participants and their families for their ongoing support to our research.

## Contributors

E.A., D.B.T, D.M. and T.R.M. conceived the study. S.K recruited the patients. E.A., M.K. and D.M. collected the data. E.A. and D.M. analysed the data. E.A., D.M. and T.R.M. wrote the manuscript with inputs from all co-authors. T.R.M. secured funding.

## Ethics Approval

The study was approved by the UK Health Research Authority (HRA) and Health and Care Research Wales (HCRW) committee (18/LO/0474).

