## Supplemental Material for "Reassessing referred sensations following peripheral deafferentation and the role of cortical reorganisation"

### EXTENDED BEHAVIOURAL METHODS

***Referred sensation task instructions***

*"In this task you will receive several kinds of stimuli from the vibrotacile stimulators placed on your body. They will feel as gentle vibrations on your skin. Some of these stimulations have been proven to stimulate a newly discovered type of nervous fibres. By doing so, they are able to evoke tactile sensations not only on the actually stimulated area of skin but also on regions of the body further away, and in particular, at the most distal extremities such as the hands or phantom hands. Indeed, due to the configuration of the peripheral nervous system, the hands are the sites at which the dual sensation most likely occurs. In this study we are interested in investigating whether there are sites on your body more responsive to this stimulation, meaning sites that when stimulated are more likely to give rise to these secondary more subtle sensations on the hands. The other types of stimulation stimulate alpha and beta fibres and convey classical information about touch to your brain, without being able to evoke other secondary sensations.*

*On each trial, you will first be cued to which kind of stimulation is coming. Thus, a red circle on the monitor indicates that in the following trial you will receive special stimulation, i.e. the "special" one capable of causing secondary subtle sensations in your hands. A grey circle will appear on the monitor if in the following trial you will receive the classical stimulation. After the coloured circle, you will receive the vibrotactile stimulus. The computer will then ask you whether you felt only one sensation (on the stimulated body site) or more than one. Please press the left pedal to respond that you only felt one and the right pedal to respond that you felt more than one. Please take your time to respond, as it usually takes a while for this interference process to happen. If you responded that you only felt one, you will start the following trial. If you responded that you felt more than one, another screen will appear asking whether you felt the secondary sensation on the left or right hand. Please respond with the left pedal if you felt it in the left or phantom hand and with the right pedal if you felt it in the right hand. The following trial will then start, and the process will be repeated."*

### EXTENDED fMRI METHODS

***Participants***

One Amputee was not able to participate in the scanning session due to MRI safety concerns and another Amputee only completed the body task due to time constraints. The proportion of participants with intact/dominant right hand was matched between Amputees and both One-handers ($X^2_{(1)}$=.942, p=.503) and Two-handers ($X^2_{(1)}$=2.670, p=.184). Amputees' gender was also matched to both One-handers ($X^2_{(1)}$=2.948, p=.106) and Two-handers ($X^2_{(1)}$=2.948, p=.106). Statistically significant differences for age were found between Amputees and One-handers ($t_{(34)}$=2.280, p=.029) and Two-handers ($t_{(37)}$=2.424, p=.020). Age covariates were therefore included when comparing between these groups.

***Functional MRI tasks***

Prior to entering the scanner room, participants were thoroughly instructed, and all movements were practiced in front of the experimenter to ensure they were performed correctly.

For the body task (used as functional localiser in [1], participants were visually instructed to move one of five body-parts: intact/dominant hand (for participants with a missing hand and Two-handers, respectively), fingers flexion and extension, residual/non-dominant arm (flexing the most distal residual joint for participants with a missing hand and the elbow for Two-handers), right or left toes (wiggling the toes) or lips (puckering the lips). An additional condition involving the missing/non-dominant hand was also included but will not be further described as it was not included in the present analyses. Movements were repeated at a constant instructed pace for a period of 12s, interleaved with 12s of rest. Each condition was repeated 4 times in a pseudo-random order.

For the face task, the full details of the procedures and acquisition parameters can be found in [2]. In short, participants were instructed to perform one of five movements: raise the eyebrows (i.e., forehead), flare nostrils (i.e., nose), puckering lips (i.e., lips), and flex the left or right thumb (or phantom thumb, if available). When phantom sensations were not present, participants with a missing hand were asked to imagine performing the movement. Note that this dataset was used to determine the relationship between the phantom hand and the face representation, and for this reason we did not include the congenital one-handers in this analysis. An additional condition involved tapping the tongue to the roof of the mouth. However, since the inner mouth was not investigated in our behavioural task, we excluded this condition from our analysis in the present study. Instructions and pace were provided visually via a screen, resulting in 5 cycles of movement per 8 seconds block. Each movement block was repeated 4 times per run, which also comprised 5 blocks of rest used as baseline.

Conditions were pseudo-randomly distributed, such that each condition was equally preceded by all other conditions. To confirm that appropriate movements were made at the instructed times, whenever possible – task performance was visually monitored online for both tasks.

***MRI data acquisition***

MRI images were acquired using a 3T Prisma MRI scanner (Siemens, Erlangen, Germany) with a 32-channel head coil. Functional data were obtained using a multiband T2*-weighted pulse sequence with a between-slice acceleration factor of 4 and no in-slice acceleration. The following acquisition parameters were used: TR = 1450 ms; TE = 35 ms; flip angle =70°; voxel size = 2 mm isotropic; imaging matrix = 106 x 106; FOV = 212 mm. 72 slices were oriented in the transversal plane covering the entire brain. Each dataset comprised one and three functional task-related block-design runs (for the body and face tasks respectively). Field-maps were acquired for field unwarping. A T1-weighted sequence (MPRAGE, TR = 2530 ms; TE = 3.34 ms; flip angle = 7°; voxel size = 1 mm isotropic) was used to obtain anatomical images.

***Functional MRI data pre-processing and analysis***

Functional data was pre-processed in FSL-FEAT (version 6.00) and included the following steps: motion correction using MCFLIRT [3]; brain extraction using BET [4]; high-pass temporal filtering with a cut-off of 280s and 119s for the body and face task respectively; and finally spatial smoothing using a Gaussian kernel with a full width at half maximum of 5mm and 3mm for the body and face task respectively. Field maps were used for distortion correction. For the face task, a midspace between the different functional runs was calculated for each participant, i.e., the average space in which the images are minimally reorientated. Each functional run was then aligned to the midspace and registered to each individual structural T1 scan using FMRIB's Linear Image Registration Tool (FLIRT), optimised using Boundary-Based Registration [5].

We focused on the S1 hand region, though marginal contribution from M1 may have affected activity profiles due to its spatial proximity. The S1 hand region of interest (ROI) was defined bilaterally for each individual on a template surface using probabilistic cytoarchitectonic maps, by selecting nodes showing maximal probability for the grey matter of Brodmann areas (BAs) 3a, 3b, 1 and 2 [6] approximately 1cm below and above the hand knob. This criterion defined a more conservative hand region than in previous research [6–8], in order to minimise overlap with the neighbouring face/arm areas. Structural T1-weighted images were used to reconstruct the pial and white-grey matter surfaces using Freesurfer. Surface co-registration across hemispheres was done using spherical alignment. The anatomical hand ROIs were projected into the individual brains via the reconstructed individual anatomical surfaces. For visualisation (Figure 2 and S1), S1 ROIs of each participant were projected to MNI152 space using the nonlinear registration carried out by FNIRT. Participant information regarding the side of missing/non-dominant hand were used to sagittal-flip data, such that the ROIs contralateral to the missing hand were always represented in the right hemisphere. ROIs of all participants were then concatenated into a single volume to produce a consistency map (i.e., how many participants have their ROIs overlapping in the MNI space). Resulting consistency maps were then projected to a group cortical surface [9] using Connectome Workbench (v1.4.2).

***Multivariate representational similarity analysis***

The dissimilarity between activity patterns within each S1 hand ROI was computed at the individual level for each pair of movements using cross-validated squared Mahalanobis distance [10]. Multidimensional noise normalisation was used to increase reliability of distance estimates (noisier voxels are down-weighted), based on the voxel's covariance matrix calculated from the GLM residuals. Due to cross-validation, the expected value of the distance is zero (or negative) if two patterns are not statistically different from each other, and significantly greater than zero if the two representational patterns are different [10]. Larger distances for movement pairs therefore suggest greater information content. The resulting representational pairwise distances between each of the facial conditions and the thumb (phantom/nondominant and intact/dominant, in Amputees and Two-handers respectively) were extracted. The analysis was conducted on an adapted version of the RSA Toolbox in MATLAB [11], customised for FSL [12].

***Additional statistical analyses***

To identify which body-parts were driving the observed remapping in the missing-hand area, independent samples t-tests were used to assess group differences between one-handed groups and Two-handers, using Bonferroni correction of alpha levels ($\alpha$=.01) to account for comparisons across the five body-parts.

### RESULTS

### FMRI DATA

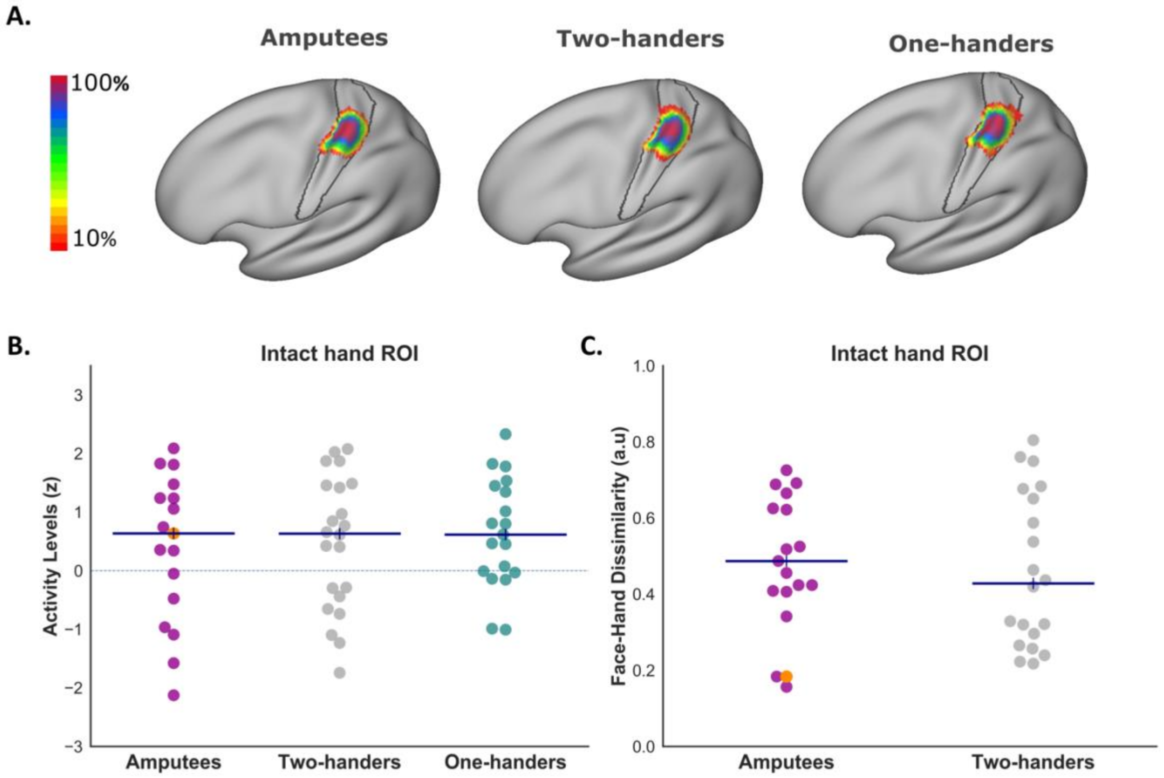


**Figure S1. fMRI results in the intact hand area. A)** Inter-participant consistency maps for the intact/dominant hand S1 regions of interest (ROIs) across the three groups. Annotations are as denoted in Figure 2A. **B)** Average BOLD activity levels in the intact/dominant hand ROI across groups. Annotations are as denoted in Figure 2C. **C)** Face-hand representational content in the intact hand area. Annotations are as denoted in Figure 3C.

#### *Body-parts undergoing S1 remapping (full-sample analysis)*

We assessed which body-parts were driving the increased activity in the missing-hand area (non-dominant in Two-handers) shown in Figure 2C, independent of individual participant's responses in the behavioural task (i.e., as reported in the main text). Since we were considering the contributions of five body-parts, we adjusted our significance (alpha) levels to 0.01. Compared to Two-handers (N=22), Amputees (N=17) exhibited increased activity for the intact Hand ($t_{(37)}$=-4.422, p<.001, d=-1.428) and decreased activity for the residual Arm ($t_{(37)}$=2.999, p=.005, d=.969). No increased activity was found in Amputees for the Lips ($t_{(37)}$=-1.664, p=.105, d=-.537, $BF_{10}$=.920). We also found a significant increase in activity for the Foot of the intact side ($t_{(37)}$=-2.978, p=.005, d=-.962) but not for the Foot of the missing side ($t_{(37)}$=-2.242, p=.031, d=-.724, $BF_{10}$=2.137). Taken together, we only find clear and consistent contribution from the intact hand to the identified group differences in activity profiles involving Amputees.

We next considered differences between the congenital One-handers and the Two-handers. Compared to Two-handers (N=22), One-handers (N=19) showed increased activity for the residual Arm ($t_{(39)}$=-2.969, p=.005, d=-.930) and the Foot of the missing side ($t_{(39)}$=-3.238, p=.002, d=-1.014). In addition, trends for increased activity were found for the Lips ($t_{(39)}$=-

2.472, p=.018, d=-.774) and the Foot of the intact side ($t_{(39)}$=-2.653, p=.011, d=-.831). No significant differences were found for the intact Hand ($t_{(39)}$=-1.384, p=.174, d=-.433, $BF_{10}$=.652). Taken together, these results indicate that residual arm is the prominent driver of observed group differences in activity levels involving One-handers.

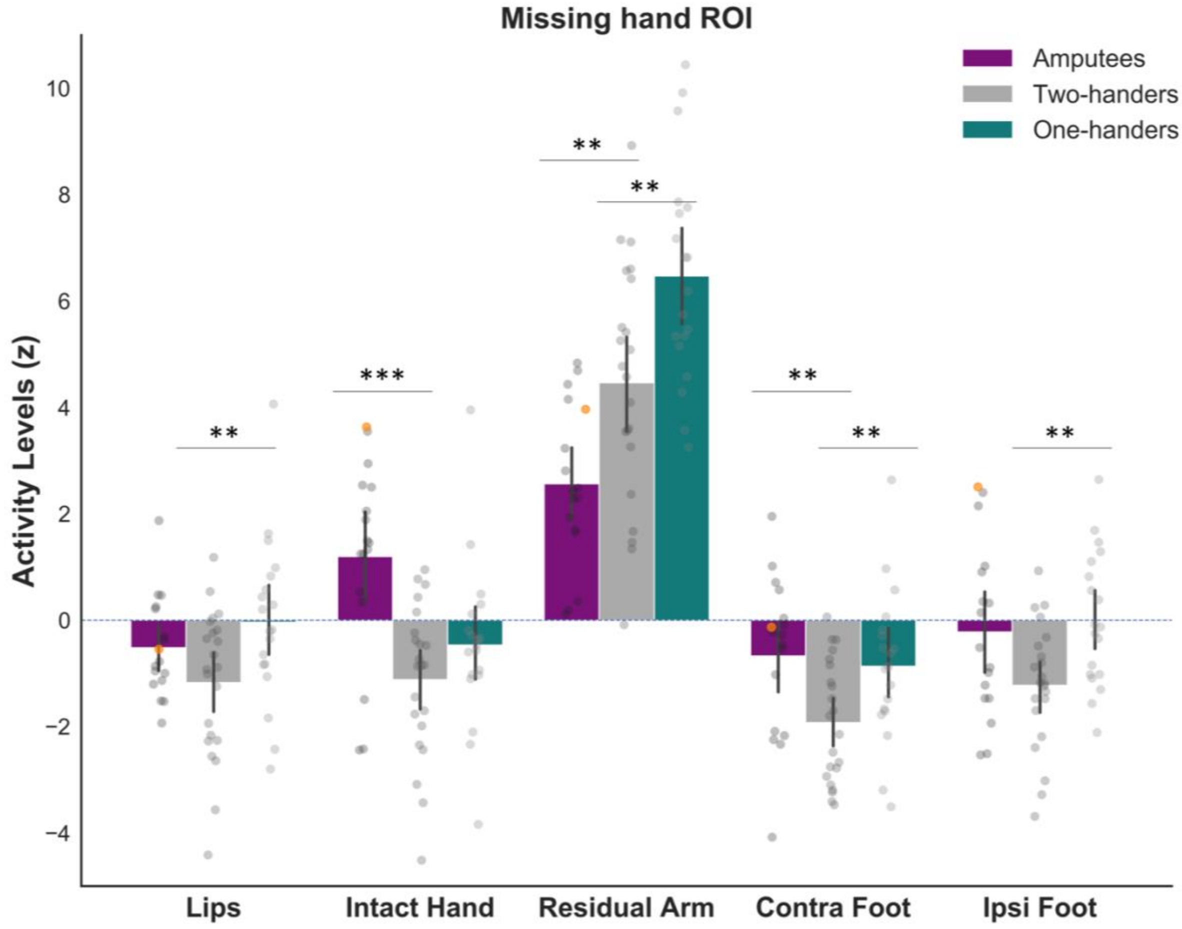


**Figure S2. Breakdown of the fMRI activity evoked by body-parts in the missing-hand area.** Average BOLD activity levels in S1 missing/non-dominant hand area, evoked in each group by movement of the Lips, Intact/Dominant hand, Residual/Non-dominant Arm, Foot Contralateral and Ipsilateral to the missing/non-dominant hand. Each grey dot represents one participant. Participant Amp05 (who reported high rates of referred sensations in the behavioural task – see Figure 1) is highlighted in orange. Asterisks indicate significant group differences: **p < 0.05, ***p < 0.001.

#### ***Sub-sample analysis based on referred sensations reports***

We considered whether we can find evidence for group differences in activity levels while specifically focussing on the sub-set of individuals in each group who reported experiencing referred sensations during the behavioural task. Compared to Two-handers (N=14), both Amputees (N=12) ($t_{(24)}$=-2.707, p=.012, $d$=-1.065) and One-handers (N=12) (U=18, p<.001, $r_B$=-.786) showed increased activity levels in the missing-hand area (i.e., remapping), resulting in a significant interaction between Groups and Hemispheres ($F_{(2,34)}$=5.604, p=.008, $\eta^2$=.045; age as covariate; non-parametric equivalent: $X^2$=11.676, p=.003). No difference in activity levels in the missing-hand area was found between Amputees and One-handers (U=82, p=.590, $r_B$=.139, $BF_{10}$=.419). No significant correlation was found between fMRI activity levels in the missing-hand area and chronic PLP in Amputees (N=12, $r_{Tau}$=.162, p=.482, $BF_{10}$=.461).

We then assessed which body-parts were driving this increased activity in the missing-hand area. As above, alpha levels were adjusted to 0.01 to correct for the five comparisons across body-parts. Compared to Two-handers (N=14), Amputees (N=12) showed increased activity levels for the intact Hand ($t_{(24)}$=-4.032, p<.001, d=-1.586) and for the Foot on the intact side (U=31, p=.005, $r_B$=-.631). A trend for increased activity was also found for the Foot on the missing side ($t_{(24)}$=-2.690, p=.013, d=-1.058, $BF_{10}$=4.274). No differences were found for the Lips ($t_{(24)}$=-1.246, p=.225, d=-.490, $BF_{10}$=.640) or the residual Arm ($t_{(24)}$=1.022, p=.317, d=.402, $BF_{10}$=.533). Compared to Two-handers (N=14), One-handers (N=12) exhibited increased activity for the residual Arm ($t_{(24)}$=-3.802, p<.001, d=-1.496) and for the Foot of the missing side ($t_{(24)}$=-2.875, p=.008, d=-1.131). No differences were found for the Lips ($t_{(24)}$=-2.285, p=.031, d=-.899, $BF_{10}$=2.245), for the Foot of the intact side (U=50, p=.085, $r_B$ =-.405, $BF_{10}$=1.067) or for the intact Hand (U=59, p=.212, $r_B$=-.298, $BF_{10}$=0.643).

Finally, we also compared the representational dissimilarity between activity patterns evoked by face and the contralateral thumb movement and we found no significant differences between Amputees and Two-handers ($F_{(1,17)}$=.176, p=.680, $\eta^2$=.009), and no interaction with the Hemisphere ($F_{(1,17)}$=.008, p=.929, $\eta^2$<.001). Follow-up comparisons revealed no significant difference between Hemispheres in Amputees ($t_{(8)}$=.316, p=.760, d=.105, $BF_{10}$=.336), as well as no group difference in the dissimilarities observed in the missing/non-dominant hand area ($t_{(17)}$=-.405, p=.690, d=-.186, $BF_{10}$=.429)).

**REFERRED SENSATION TASK**

***Referred sensations evoked non-facial body-parts***

While referred sensations were originally reported to be evoked by the face and residual arm on the phantom hand [13–16], later reports also included multiple body-parts, including the intact hand and arm, and the feet [13,17–21]. Here we examined whether participants across groups tended to report more referred sensations on their phantom/missing/non-dominant relative to their intact/dominant hand following stimulation of non-facial sites (i.e. (Intact/Dominant Arm, Intact/Dominant Hand, Foot Ipsilateral and Contralateral to missing/non-dominant hand – 48 trials). Note that here the Residual Arm site was excluded, due to its distinct potential mechanism for inducing referred sensations (via peripheral reinnervation, see Introduction). We found that non-face evoked referred sensations were not reported more frequently on the phantom/missing/non-dominant rather than on the intact/dominant hand in Amputees (N=12) (Z=53.5, p=.265, $r_B$=.372, $BF_{10}$=.519), One-handers (N=7) (Z=7.5, p=.310, $r_B$=-.464, $BF_{10}$=.499), and Two-handers (N=9) (Z=25.5, p=.326, $r_B$=.417, $BF_{10}$=.647). Moreover, no significant group difference emerged ($X^2$=1.822, p=.402), with Amputees' phantom lateralised non-face evoked referred sensations not significantly different from One-Handers' (U=56, p=.249, $r_B$=.333, $BF_{10}$=.666) or Two-handers' (U=51.5, p=.885, $r_B$=-.046, $BF_{10}$=.415), and no significant differences between One-handers and Two-handers (U=42.5, p=.262, $r_B$=.349, $BF_{10}$=.669).

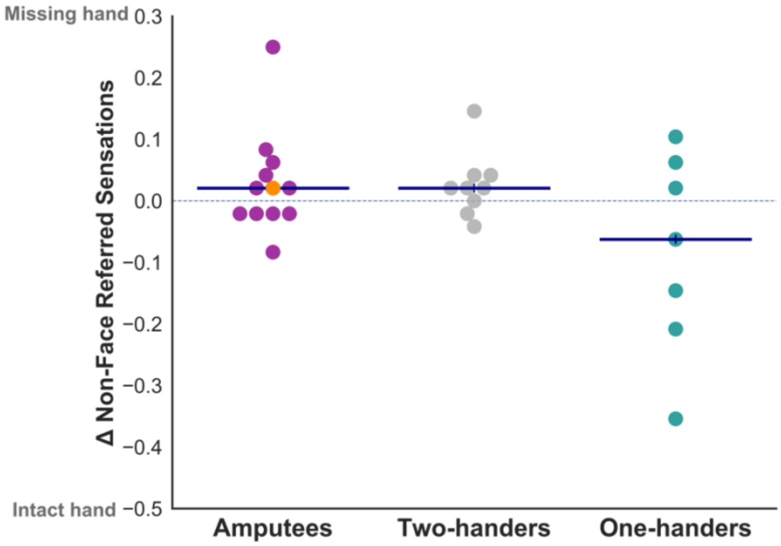


**Figure S3. Lateralised referred sensations across the non-facial stimulation sites**. Scores are calculated by subtracting the proportion of referred sensations reported on the intact/dominant hand from the proportion of responses on the phantom/missing/non-dominant hand, in Amputees, One-handers and Two-handers, respectively. Participants reporting zero referred sensations across these 48 trials are excluded (52% of total sample). Each dot represents one participant, horizontal blue lines represent group medians. Participants Amp05 is highlighted in orange.
